# The fungal pathogen and mycobiota diversity in respiratory samples from children with cystic fibrosis

**DOI:** 10.1101/2023.03.16.23287307

**Authors:** Rebecca Weiser, Katherine Ronchetti, Jo-Dee Tame, Sven Hoehn, Tomasz P. Jurkowski, Eshwar Mahenthiralingam, Julian T. Forton

**Author notes:** **Corresponding author at:** Microbiomes, Microbes and Informatics Group, Organisms and Environment Division, School of Biosciences, Cardiff University, Sir Martin Evans Building, Park Place, Cardiff, UK.

## Abstract

**Background:** The prevalence of fungi in cystic fibrosis (CF) lung infections is poorly understood and studies have focused on adult patients. We investigated the fungal diversity in children with CF using brochoalveolar lavage (BAL) and induced sputum (IS) samples to capture multiple lung niches.

**Methods:** Sequencing of the fungal ITS2 region and molecular mycobiota diversity analysis was performed on 25 matched sets of BAL-IS samples from 23 children collected as part of the CF-SpIT study (UKCRN14615; ISRCTNR12473810).

**Results:** *Aspergillus* and *Candida* were detected in all samples and were the most abundant and prevalent genera, followed by *Dipodascus, Lecanicillium* and *Simplicillium*. The presumptive CF pathogens *Exophiala, Lomentospora* and *Scedosporium* were identified at variable abundances in 100%, 64%, and 24% of sample sets, respectively. Fungal pathogens observed at high relative abundance (≥40%) were not accurately diagnosed by routine culture microbiology in over 50% of the cohort. The fungal communities captured by BAL and IS samples were similar in diversity and composition, with exception to *C. albicans* being significantly increased in IS samples. The respiratory mycobiota varied greatly between individuals, with only 13 of 25 sample sets containing a dominant fungal taxon. In 11/25 BAL sample sets, airway compartmentalisation was observed with diverse mycobiota detected from different lobes of the lung.

**Conclusions:** The paediatric mycobiota is diverse, complex and inadequately diagnosed by conventional microbiology. Overlapping fungal communities were identified in BAL and IS samples, showing that IS can capture fungal genera associated with the lower airway. Compartmentalisation of the lower airway presents difficulties for consistent mycobiota sampling.

**What is already known on this topic:**

- Fungal lung infections in people with CF are poorly described and reports are largely based on conventional culture data from adults, with limited studies on the role of fungal infections in children.

**What this study adds:**

- We uniquely used culture-independent analysis to interrogate the mycobiota in different respiratory samples (bronchoalveolar lavage and induced sputum) from 23 children with CF aged between 1-18 years.
- *Aspergillus* and *Candida* were detected in 100% of samples at varying levels, and whilst high relative abundances (>50%) of these genera and the emerging *Exophiala* were detected in multiple samples from children over the age of 12, culture-based diagnostics failed to accurately identify them.
- There were differences in the fungal communities in different regions of the lung from the same individual, suggesting mycobiota compartmentalisation.

**How this study might affect research, practice or policy:**

- Fungal prevalence rates in children with CF are underestimated and inaccurately diagnosed by routine culture, which reflects a need for longitudinal respiratory sampling, molecular analyses and improvement in conventional mycology diagnostic practice.

## 1. INTRODUCTION

The respiratory tract in people with cystic fibrosis (pwCF) harbours a complex polymicrobial community of bacteria, fungi and viruses. The bacterial diversity in CF lung infections has been widely examined by both conventional culture-based approaches and culture-independent microbiota studies (1). A variety of bacterial groups have been identified as consistently associated with the CF lung infections, including those considered ‘traditional’ pathogens such as *Pseudomonas* and *Staphylococcus*, and those whose role in disease progression is still unclear, such as the anaerobes *Prevotell*a and *Veillonella* (1).

In contrast, the prevalence, diversity and clinical epidemiology of fungi in CF lung infections in children is poorly understood. Historically, fungi were often disregarded when detected by culture as either contaminants or harmless colonisers of the oral cavity and respiratory tract (2). In addition, conventional sampling may underestimate the fungal burden in CF, since fungal culture can be highly variable and incapable of detecting the full spectrum of fungal diversity (2). Mycobiota studies, based on culture-independent analysis of the fungal taxa present, are emerging but lag behind bacterial diversity studies. In part, this is due to the comparatively limited tools and databases available for fungal DNA sequence analysis and taxonomic classification (3, 4).

The filamentous fungus *Aspergillus fumigatus* and the yeast *Candida albicans* are the most common fungal species encountered as a respiratory infection and can be identified in over 50% of pwCF using conventional mycology approaches (5). Other *Aspergillus species* (*A. niger, A. flavus, A. nidulans* and *A. terrus*) and *Candida* species (*C. parapsilosis, C. glabrata* and *C. krusei*) have also been detected as well as *Scedosporium* species, *Lomentospora prolificans* and *Exophiala dermatitidis* (6). The clinical consequences of *A. fumigatus* in pwCF have been most defined (including allergic, bronchitic and invasive disease), whilst the other fungal groups mentioned are presumed potential pathogens and an increasing number of studies have documented their detection and disease associations (5-7).

The majority of fungal lung infection research in CF has focused on adults and we currently know very little about the fungi in the airways of children with CF, with the exception that fungal colonisation increases with age (5). Long-term population data for fungal infection in pwCF is limited. The UK CF Registry for example has only been tracking the rate of *Aspergillus* lung infection since 2017. In 2021, *Aspergillus* prevalence in the UK CF population was 6.1% in children age <16 years and 10.3% in adults; no other fungal pathogens are currently reported in the Registry (8).

In this study we apply molecular mycobiota analysis to respiratory samples collected by the Cystic Fibrosis Sputum Induction Trial (CF-SpIT) (9, 10) to explore the fungal diversity in children with CF. By using both bronchoalveolar lavage (BAL) and induced sputum (IS) samples, we capture and describe fungal communities in the upper and lower airways.

## 2. METHODS

### 2.1 Respiratory sample collection, microbiological culture and DNA extraction

Details of the CF-SpIT study design and participants can be found elsewhere (9, 10). Sputum induction and BAL procedures were performed as previously described (9). Briefly, BAL fluid from the right middle lobe was collected and labelled as BAL1, from the left lingula as BAL2, and from the combination of right upper lobe, right lower lobe, left upper lobe and left lower lobe as BAL3. All samples were divided immediately into two aliquots. One aliquot was processed for bacteria and fungi in the microbiology laboratory of the University Hospital of Wales. The other aliquot was frozen at – 80 °C within 30 min of collection for DNA extraction. DNA extraction was performed as described previously (10). Sample sets were identified with an ID number beginning with ‘CF’ to anonymise the identity of study subjects.

### 2.2 Quantitative PCR (qPCR)

Quantification of total fungal load was performed using a TaqMan® qPCR assay targeting the fungal 18S rRNA gene as described by Liu *et al*. (2012) (11). See Supplementary Materials for further details.

### 2.3 ITS2 region sequencing and bioinformatics analysis using QIIME2

Twenty-five matched sets of BAL1-BAL2-BAL3-IS samples were subjected to ITS2 sequencing analysis. Amplification and sequencing of the ITS2 region was performed by Novogene, generating 250 bp paired-end reads using Illumina sequencing technology. The primers used for amplification of the ITS2 region were ITS3-2024F (5’-GCATCGATGAAGAACGCAGC-3’) and ITS4-2409R (5’-TCCTCCGCTTATTGATATGC-3’) (12). The sequencing run contained a mock fungal community (Mycobiome Genomic DNA Mix, MSA-1010; ATCC) and 3 DNA extraction controls. Along with the qPCR data, these controls were used to confirm accurate mycobiota profiling and determine subsampling thresholds in downstream analyses (further details in Supplementary Materials).

Bioinformatic analysis was carried out using a virtual machine hosted by the Cloud Infrastructure for Microbial Bioinformatics (CLIMB) consortium (13). Quality control and Illumina adapter trimming of the raw sequencing reads was performed using FastQC v0.11.5 and Trim Galore! v0.4.3 for paired end reads. Trimmed reads were imported into QIIME2 (14) (q2cli version 2020.11.1), the q2-ITSxpress plugin (15) used to identify and trim the ITS2 region sequences and the DADA2 plugin (16) used to group sequences into amplicon sequence variants (ASVs). Taxonomic classification of fungal ITS2 ASVs was performed using the q2-feature-classifier (17), with the classifier first being trained using the UNITE database v8.2 dynamic dataset (18). Bioinformatics scripts are available at https://github.com/Beky-Weiser/CFSpIT-ITS2-Microbiota-Analysis.

### 2.4 Fungal diversity and statistical analyses

The fungal ASV table, fungal taxonomy and sample metadata files were imported into R statistical software v4.0.3 (19) using the package phyloseq v1.34.0 (20). Low abundance ASVs representing less than 0.01% of the total number of sequence reads were removed. Subsampling to 6000 sequence reads was performed and coverage for all 100 samples was calculated at ≥99.5%. See Supplementary Materials for further details.

Further data handling and statistical analyses were carried out in R statistical software and Microsoft Excel. Alpha and beta diversity indices were calculated using the R package vegan (version 2.6-2) (21). Statistical differences in alpha-diversity (Shannon index) and sequence read proportions were evaluated using the Friedman test for non-parametric dependent data with patient as the blocking factor, and *post hoc* Conover pairwise comparisons with Bonferroni-adjusted P values (R package PMCMR v4.4). Beta diversity was analysed using the Bray-Curtis dissimilarity measure and non-metric multidimensional scaling (NMDS) ordination performed to visualise sample distributions in two dimensions. Differences between sample groups were analysed using Permutational ANOVA (PERMANOVA; adonis function in vegan package). Differences between genus relative abundance in sample types was determined by GAMLESS-BEZI as previously described (22). Indicator species analysis was performed using the R-package indicspecies v1.7.12 (23) to identify ASVs that were significantly associated with sample types. The relationship between Shannon diversity and age was investigated using linear regression and linear mixed models with patient as the random effect (R package nlme). Further details and R code are available at https://github.com/Beky-Weiser/CFSpIT-ITS2-Microbiota-Analysis.

### 2.5 Data availability

Raw sequence data have been submitted to the European Nucleotide Archive under project number PRJEB60511.

## 3. RESULTS

### 3.1 Patient sample sets and mycobiota analysis overview

Mycobiota profiles were obtained for 25 matched BAL1-BAL2-BAL3-IS sets from 23 children with CF aged between 1.1 and 17.7 years. Overall, 668 unique ASVs were identified across the 100 samples, of which 442 could be classified at genus level (130 unique genera) and 321 could be classified at species level (159 unique species). Strikingly, 143 ASVs could not be classified beyond ‘kingdom Fungi’ and remained unidentified. Detailed sequencing statistics are given in Supplementary Materials.

In relation to CF fungal genera of particular interest, at a presence/absence level mycobiota analysis detected *Aspergillus* and *Candida* in 100% of sample sets and the other presumptive pathogens *Exophiala, Lomentospora* and *Scedosporium* in 100%, 64%, and 24% of sample sets, respectively (See Supplementary Materials).

### 3.2 Mycobiota analysis was superior to culture for fungal detection and identification

Culture data was available for all 100 samples belonging to the 25 matched BAL1-BAL2-BAL3-IS sets. Although mycobiota analysis identified the presence of fungi in 100% of samples, routine culture only identified fungi in 20% (Supplementary Table S1) and fungal growth was indicated broadly as “*Candida*”, “*Aspergillus*”, “yeast-like growth” or “fungal mycelial-like growth”. In addition, culture-based diagnostics failed to identify fungal genera that were at high relative abundance in mycobiota analysis as follows: *Candida* was identified between 43.5% and 98.2% relative abundance in 10 individuals, but only 5 were reported as culture positive; two individuals had 83.4% and 96.6% relative abundance of *Aspergillus*, with one case correctly identified, but the other recorded as *Candida* culture positive; and *Exophiala* was seen at 62.9% and 80.1% relative abundance in two individuals, and in both instances *Aspergillus*/*Candida* was reported from culture (Table S1).

### 3.3 Fungal communities in BAL and IS samples were similar in diversity and composition

The alpha diversity (within-sample diversity) was similar across all sample types (Figure 1, panel A and B) and BAL1, BAL2, BAL3 and IS samples also overlapped in their fungal community composition (beta diversity; Figure 1, panel C). The top 25 most abundant ASVs across the dataset represented a similar proportion of total sequence reads for BAL1, BAL2, BAL3 and IS (Figure 1, panel D) and these proportions were not significantly different.

**Figure 1.**
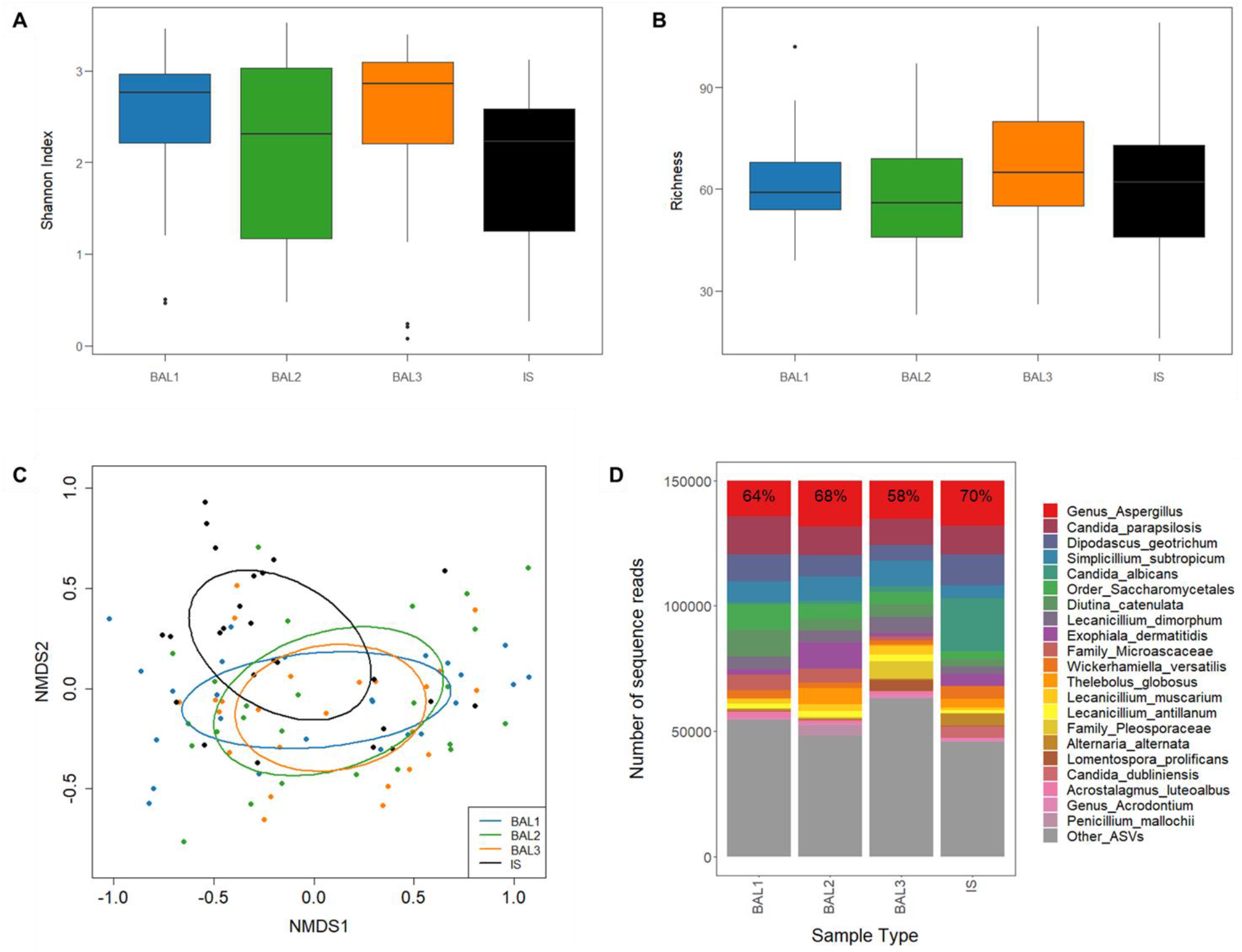
Fungal communities in BAL and IS samples were similar in diversity and composition. Boxplots **(A, B)** indicate the spread of the alpha diversity values across sample types. No significant differences were found for the Shannon Index **(A)** or ASV richness **(B)**. NMDS ordination of Bray-Curtis dissimilarity distances **(C)** indicated that BAL samples clustered together and IS samples overlapped in composition with BAL samples; there were no significant differences between the groupings. The top 25 most abundant ASVs represented a similar proportion of the total sequence reads in BAL1 (64%), BAL2 (68%), BAL3 (58%) and IS (70%) samples **(D)**. In **(D)** the top 25 most abundant ASVs have been consolidated to lowest taxonomic rank and their cumulative percentage indicated at the top of the stacked bar charts, all other ASVs have been consolidated and are shown in grey. The taxa colour key is displayed to the right of the figure.

### 3.4 Key differences between BAL and IS samples were linked to *Candida* species

In ranked order, the most abundant and prevalent fungal genera identified across the entire dataset were *Candida, Aspergillus, Dipodascus, Lecanicillium* and *Simplicillium*, and this trend was largely mirrored when sample types were considered separately (Table 1). Notably, *Candida, Aspergillus* and *Simplicillium* were identified in 100% of samples although their relative abundances were variable (<1% to >99%; Supplementary Table S2). *Exophiala, Lomentospora* and *Scedosporium* were also detected in BAL and IS samples, ranking 7^th^ (83% prevalence), 17^th^ (41% prevalence) and 89^th^ (11% prevalence) most abundant across all samples, respectively. The relative abundances of *Aspergillus, Dipodascus, Lecanicillium, Simplicillium, Exophiala, Lomentospora* and *Scedosporium* were not significantly different between the sample types, but *Candida* had significantly increased relative abundance in IS compared to BAL1 (p=0.03), BAL2 (p=0.001) and BAL3 (p=0.006) samples (Supplementary Table S2).

**Table 1.** The most abundant and prevalent fungal genera across the different sample types

| Rank abundance | BAL123-IS | BAL1 | BAL2 | BAL3 | IS |
| --- | --- | --- | --- | --- | --- |
| 1 | *Candida*<br>(100) | *Candida*<br>(100) | *Aspergillus*<br>(100) | *Aspergillus*<br>(100) | *Candida*<br>(100) |
| 2 | *Aspergillus*<br>(100) | *Aspergillus*<br>(100) | *Candida*<br>(100) | *Lecanicillium*<br>(100) | *Aspergillus*<br>(100) |
| 3 | *Dipodascus*<br>(88) | *Dipodascus*<br>(84) | *Simplicillium*<br>(100) | *Candida*<br>(100) | *Dipodascus*<br>(96) |
| 4 | *Lecanicillium*<br>(98) | *Diutina*<br>(76) | *Lecanicillium*<br>(100) | *Simplicillium*<br>(100) | *Lecanicillium*<br>(92) |
| 5 | *Simplicillium*<br>(100) | *Lecanicillium*<br>(100) | *Exophiala*<br>(84) | *Dipodascus*<br>(88) | *Simplicillium*<br>(100) |

ASVs (n=668) were consolidated to genus and genera ranked by total read number (abundance). Percentage prevalence (samples positive for a particular genus) is given in brackets under genus ID.

At species level, the total numbers and proportions of sequence reads assigned to different species within a genus were similar, except for *Candida* (Figure 2; Supplementary Figure S1). Eight *Candida* species were identified across the dataset, of which *C. parapsilosis, C. albicans, C. krusei* and *C. dubliniensis* were the most abundant. The total number of reads assigned to *C. parapsilosis* were similar for BAL1, BAL2, BAL3 and IS, but reads were increased for *C. albicans* and *C. dubliniensis* in IS and for *C. krusei* in BAL1. This led to differences in *Candida* species sequence read proportions between BAL and IS samples (Figure 2). Indicator species analysis (Supplementary Table S3) revealed significant associations between two *C. albicans* ASVs and IS samples (ASV10, p=0.02; ASV14, p=0.03) and one *C. krusei* ASV (ASV29, p=0.008) and BAL1 samples.

**Figure 2.**
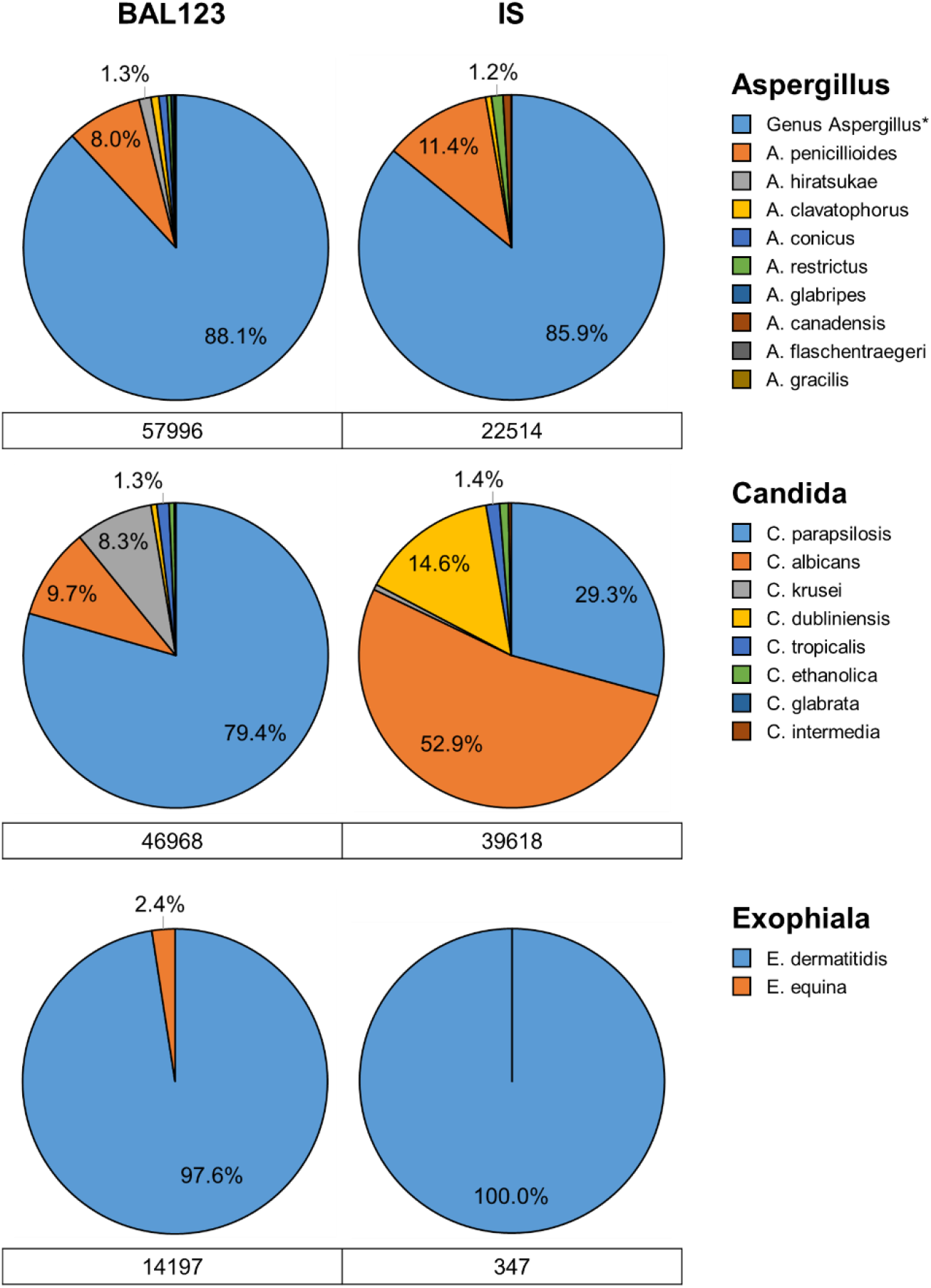
Distribution of sequence reads at species level in BAL and IS samples. The two most abundant and prevalent genera (*Candida* and *Aspergillus*) and presumptive CF pathogen *Exophiala* are shown. Reads from BAL1, BAL2 and BAL3 samples are consolidated into one group as ‘BAL123’. Pie charts display the proportion of sequence reads assigned to different species within a genus, proportions ≥1% are displayed. Total sequence reads for each genus are below each pie chart. *Genus_Aspergillus comprised 21 ASVs, 4 of which (56% of the Genus_Aspergillus sequence reads) were further classified to *A. fumigatus* and were present in 98% of samples (see Supplementary Materials).

### 3.5 There was large variation between fungal communities in samples from different individuals

A large number and variety of fungal taxa were identified across the 100 respiratory samples and each individual possessed a unique respiratory mycobiota. Examples of different profile types are given in Figure 3 and the full set is displayed in Supplementary Figure S2. On average, each sample comprised 62 unique ASVs (range: 16-109) and the 25 most abundant ASVs in each sample averaged a cumulative relative abundance of only 83.6% (range: 13.0-99.7%).

**Figure 3.**
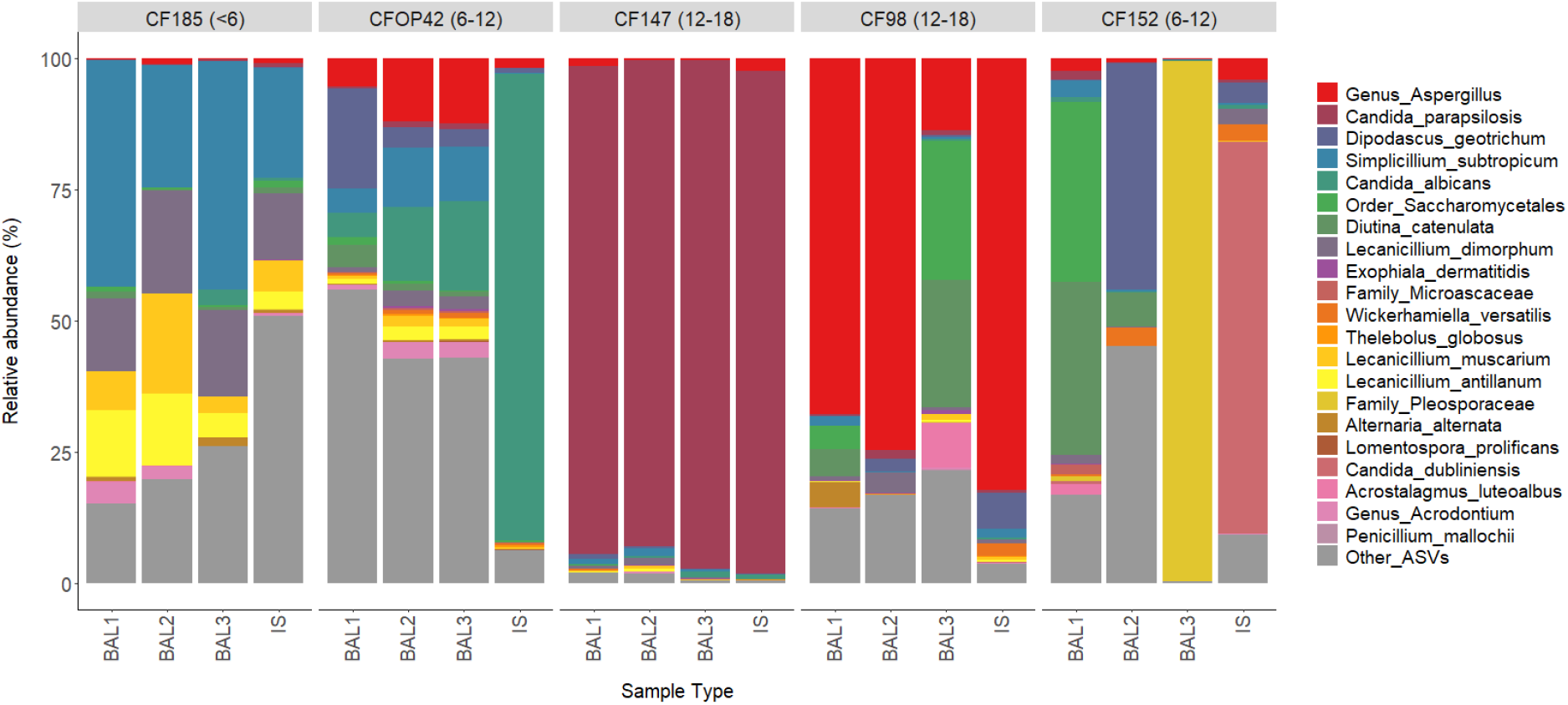
BAL1, BAL2, BAL3 and IS samples sets illustrating different mycobiota profile types. Stacked bar charts of mycobiota profiles are shown for 5 sample sets to provide examples of samples with a diverse mycobiota **(A – all samples, B – BAL samples)**, samples dominated by a single taxon **(B – IS sample, C – all samples)**, sample sets that are largely similar **(A, C)**, sample sets where one sample is dissimilar from the others **(B – IS sample, C – BAL3 sample)** and sample sets where all samples are dissimilar **(E)**. The top 25 most abundant ASVs consolidated to lowest taxonomic rank (coloured, not grey) and all other ASVs consolidated (coloured grey) are shown. The taxa colour key is displayed to the right of the figure.

The top 25 ASVs across the dataset were consolidated to their lowest taxonomic rank for further analysis. Twenty-seven out of one hundred samples belonging to 13 sample sets had a dominant fungal taxon (defined as >50% relative abundance) with the remaining 73/100 samples considered to have diverse mycobiota profiles. A higher percentage of IS samples (10/25; 40%) had a dominant fungal taxon than BAL samples (17/75; 24%). Dominant fungal taxa included *Aspergillus* (6 samples from 4 sample sets), *C. parapsilosis* (6 samples from 2 sample sets), *C. albicans* (3 samples from 3 sample sets), *E. dermatitidis* (3 samples from 2 sample sets), *Microascaceae* (2 samples from 1 sample set), *Thelebolus globosus* (2 samples from 2 sample sets) and *Pleosporaceae, Alternaria alternata, Lomentospora prolificans, C. dubliniensis* and *Penicillium mallochii* (1 sample each from 5 different sample sets) (Supplementary Figure S2).

Increased relative abundances of *Aspergillus, Exophiala* and *C. parapsilosis* were seen in both BAL and IS samples with increasing age (Supplementary Figure S3, Panel B). A trend for decreasing alpha diversity (Shannon diversity) with increasing age was observed for both BAL and IS samples, although this correlation was not significant in the cohort examined (Supplementary Figure S3, Panel A).

### 3.6 Compartmentalisation of the lower airway mycobiota can occur

BAL1, BAL2 and BAL3 directly sample the lower respiratory tract but fungal communities were not always uniform across BAL samples from the same patient (Supplementary Figure S2). To investigate this further, Bray-Curtis dissimilarity distances between BAL samples within- and between-patients were compared. The median values of the within-(0.76) and between-patient (0.88) comparisons were similar and the interquartile ranges overlapped (Figure 4), indicating that within-patient comparisons could be as dissimilar as between-patient comparisons; 11/25 sample sets fell above the 25% percentile of the between-patient comparison dataset (Figure 4).

**Figure 4.**
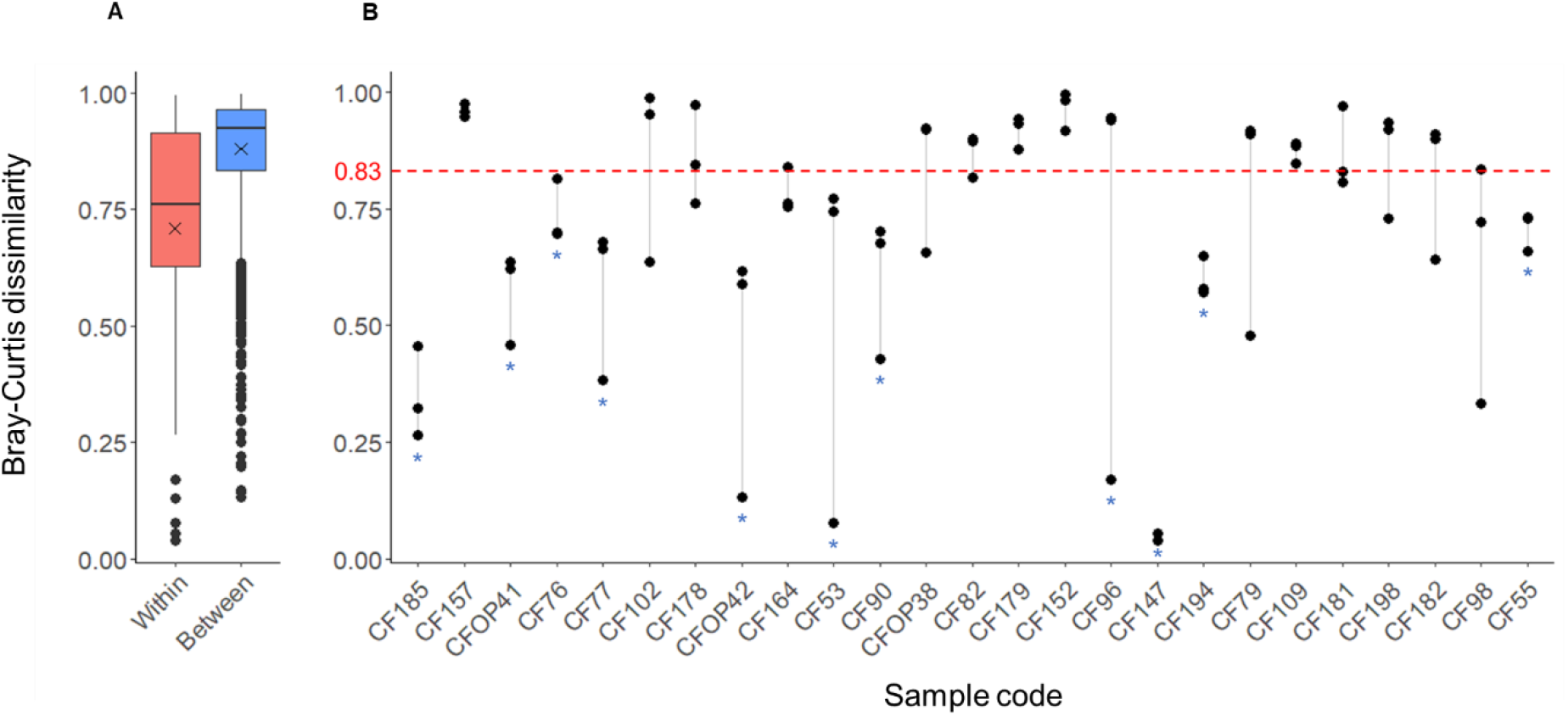
Comparison of within-patient and between-patient mycobiota diversity of BAL samples. Bray-Curtis dissimilarity distances for within-patient and between-patient comparisons are shown as follows. **(A)** Boxplots of BAL within-patient comparisons (median = 0.76 (IQR 0.63–0.91) and between-patient comparisons (median = 0.93 (IQR 0.83–0.96). The mean values are shown as a cross inside the boxplot (within = 0.71; between = 0.88). **(B)** Bray-Curtis dissimilarity distances between BAL samples taken from individual patients are visualised using strip charts; three values are plotted per patient (BAL1-BAL2, BAL1-BAL3 and BAL2-BAL3 distances), points are joined to highlight the spread. The first quartile of the between-patient data (0.83) has been highlighted by a red dotted line and 14 sets of within-patient comparisons falling below this line highlighted with a blue asterisk. The remaining 11 sets are considered highly dissimilar. Sample sets are arranged in order of increasing patient age.

At a presence/absence level only 66% (range: 20-100%) of the top 10 and 44% (range: 16-92%) of the top 25 ASVs were shared across all 3 BAL samples within a set, suggesting that the lower airway mycobiota can be compartmentalised with different fungal communities present in different lobes of the lung.

## 4. DISCUSSION

The current understanding of the epidemiology and pathogenesis of fungal lung infections in adults with CF is limited, with even less known in children with CF. In this study, we used respiratory samples collected as part of the CF-SpIT trial (9, 10) to characterise the fungal communities in the airways of children with CF between 1.1 and 17.7 years of age.

A diverse range of fungi were identified and there was large variation between different individuals. The fungal communities in BAL and IS samples overlapped in diversity and composition suggesting that IS is capable of capturing the upper and lower airway mycobiota. A key difference between the two sampling methods was the increased abundance of *C. albicans* in IS samples. This increased *Candida* signature may be linked to the nature of IS sampling, where samples pass through the upper respiratory tract and oral cavity before collection. *C. albicans* has been found in the oral mycobiota of pwCF at higher prevalence than people without CF (24).

*Aspergillus* and *Candida* species were the most prevalent fungal genera in our cohort, with both genera detected in all respiratory samples from all individuals at variable relative abundances (<1% to >99%). In addition, at a presence/absence level, the emerging fungal pathogens *Exophiala, Lomentospora* and *Scedosporium* were detected in 100%, 64% 24% of sample sets, respectively. These molecular-detection based prevalences were far higher than matched fungal culture results, and are higher than reported prevalences from other culture-based studies in both adults (5-7) and children with CF (5, 25, 26). Of all the fungal pathogens, *A. fumigatus* epidemiology has been best documented in children with CF and has reported culture-based prevalence of 2-15% in children under 6 years (5, 25, 26) and 25-36% in children between 6-18 years (5, 25). Data from the German CF registry in 2017 also identified the culture-based prevalences of other fungal groups in children as lower than in our study with *C. albicans* at 10-40%, other *Candida* species at 10-30% and *Aspergillus* (non-*A. fumigatus*), *Scedosporium* and *Exophiala* species each at less than 10% prevalence (5). Culture-independent mycobiota studies in adults have shown prevalence rates are underestimated by conventional fungal culture (27) and in this study we demonstrate the same situation in CF children.

We identified a large range of fungi including known and potential emerging pathogens, but the clinical implications of these fungi remain unclear. As highlighted in a previous study into the bacterial diversity of the samples analysed here, consideration of what level of microbiota relative abundance clinically constitutes an infection is a challenge (10), and setting a threshold for mycobiota analysis is similarly difficult. Interestingly, when an arbritrary relative abundance level of ≥40% for the presence of key fungal genera was applied to our dataset, the identification obtained by conventional microbiology was inaccurate for 8 of 14 individuals (57%; Supplementary Table S1). Given the known clinical problems associated *Aspergillus* (25, 26) and the lung function decline being seen in *Exophiala* positive CF adults within the same geographic region (28), further review of optimal fungal microbiology practices in CF is necessary.

It is also important to consider whether fungi detected in CF airways are transient or persistent colonisers, as fungi may be regularly acquired via inhalation or potentially via seeding from the oral cavity (29). These mechanisms are particularly relevant for *Aspergillus*, as it is estimated that thousands of conidia may be inhaled per day (30), and for *Candida*, as the most prevalent genus in the oral mycobiota (29). A recent mycobiota study in CF adults with longitudinal samples suggested that fungal communities were predominately composed of transient members, although there were exceptions where fungal pathogens were repeatedly isolated from the same patient (29). In our study, two patients provided sample sets of two separate occasions and whilst their profiles were not identical, fungal taxa were shared (Supplementary Figure S2; CFOP38 and CF194 shared *C. parapsilosis* and *S. subtropicum*, CF96 and CF198 shared *C. parapsilosis* and *Aspergillus*).

As the remainder of our dataset was cross-sectional, we were unable to further investigate the transience of fungal taxa, but we can speculate about the implications of differences in fungal diversity. Numerous studies have linked a decrease in bacterial diversity to a decline in lung function in CF (31) and there is evidence to suggest a similar link to fungal diversity (32). The majority of samples in our study had a diverse mycobiota but 27/100 samples belonging to 13 samples sets had a dominant fungal taxon. All 27 samples were from children over 6 years of age and there were samples sets where multiple matched samples had the same dominant fungal taxon. In these cases, the dominant fungi identified below family level were *C. parapsilosis* (CF96, CF147), *Aspergillus* (CF98) and *E. dermatitidis* (CF181), all of which have been found to dominate sputum mycobiota profiles in CF adults (27). We propose that the discovery of these presumptive fungal pathogens at high levels in multiple airway compartments for our paediatric CF cohort represents more than transient colonisation or sample contamination.

In addition to fungi previously associated with CF, we also unexpectedly discovered the predominantly environmental fungi *Simplicillium, Lecanicillium* and *Dipodascus* in 100%, 98% and 88% of all samples, respectively. Although at high prevalence, these genera had an average relative abundance of <10% and reached maximum relative abundances of 47-54%. Human infections with these genera are rare, but *Simplicillium* has been found in the pulmonary mycobiota of non-CF bronchiectasis (33) and both *Simplicillium* and *Dipodascus* have been associated with the human digestive tract (34, 35). *Lecanicillium* has been recovered from CF nebulisers (36) suggesting a potential route for acquisition of this fungus. The clinical impact of these genera in the CF respiratory mycobiota is unknown, but their detection herein warrants further correlations to be made via longitudinal study.

We previously investigated all of the samples in this study by culture-independent microbiota analysis to determine the bacterial diversity (10). Similarly, we found that the bacterial communities captured by BAL and IS samples overlapped in composition, and that in certain patients the bacterial microbiota was compartmentalised in the lower airways (10). Compartmentalisation has been found in adults and children with CF in relation to bacterial infection, but to our knowledge this is the first study reporting that different lobes of the CF lung can harbour different fungal communities. Currently, relatively few CF mycobiota studies are available, and those published have generally relied on sputum rather than targeted BAL sampling (4). There are also still numerous challenges to overcome in studying the lung mycobiota, including the difficulties of fungal DNA extraction and the lack of tools and databases for fungal DNA sequence analysis (3, 4). We experienced difficulties in fungal sequence classification, requiring the development of our own analysis strategy (see Supplementary Materials). In particular, large numbers of ASVs could not be assigned to genus level and ASV assignments needed to be scrutinised during analysis due to fungal taxonomy and nomenclature being a rapidly evolving field (37).

An important first step in our understanding the CF lung mycobiota is identifying which fungi are present, as we have done here for different respiratory sample types from children with CF. We have shown that the airway mycobiota is diverse and complex, and that presumptive CF fungal pathogens may be detected in children, with their dominance in samples potentially increasing with age. The clinical implications of fungal detection are still not understood and would require longitudinal sampling and correlations to clinical parameters to comprehensively investigate. In addition, although it was not within the scope of this study, with both bacterial and fungal diversity datasets now available for CF-SpIT samples we can begin to investigate inter-kingdom networks in children with CF. This will ultimately broaden our ecological understanding of the CF lung mycobiota as in other emerging research for adults with CF (38, 39). Finally, the overlapping and correlating mycobiota signals obtained from BAL and IS, also indicate that IS sampling can be taken forward as a straightforward means to capture CF fungal epidemiology in future studies.

## Supporting information

Supplemental Materials - ASV table and sample metadata

Supplemental Materials - methods and results

## Data Availability

Sequence data are available online at the European Nucleotide Archive under project number PRJEB60511

https://www.ebi.ac.uk/ena/browser/view/PRJEB60511

https://github.com/Beky-Weiser/CFSpIT-ITS2-Mycobiota-Analysis

## Acknowledgements

This work was funded by a research project award from the US Cystic Fibrosis foundation grant MAHENT20G0, with additional support from the Health and Care Research Wales-Academic Health Science Collaboration and Wellcome Trust Institutional Strategic Support Fund (Cardiff University).

## Author contributions

The CReDIT contributor roles taxonomy was used to recognise author contributions as follows:

Conceptualization: RW, EM, & JTF

Methodology: RW, KR, JT, SH, TPJ, EM & JTF

Software: RW, SH, & TPJ

Validation: RW, SH, TPJ, EM & JTF

Formal analysis: RW, SH, TPJ, & JTF

Investigation: RW, KR, JT, SH, & JTF,

Resources: TPJ, EM, & JTF

Data Curation: RW, SH, & JTF

Writing – original draft preparation: RW, EM, & JTF

Writing – review and editing: RW, EM, & JTF

Visualization: RW, & JTF

Supervision: TPJ, EM, & JTF

Project administration: RW, KR, JT, TPJ, EM, & JTF Funding acquisition: RW, EM, & JTF

