## Supplemental Materials - methods and results for "The fungal pathogen and mycobiota diversity in respiratory samples from children with cystic fibrosis"

**AFFLIATIONS:**

[1]Microbiomes, Microbes and Informatics Group, Organisms and Environment Division, School of Biosciences, Cardiff University, Sir Martin Evans Building, Park Place, Cardiff, UK

[2]Department of Paediatric Respiratory Medicine, Noah's Ark Children's Hospital for Wales, Cardiff, UK

[3]Department of Paediatric Physiotherapy, Noah's Ark Children's Hospital for Wales, Cardiff, UK

[4]School of Healthcare Sciences, Cardiff University, UK

[5]Molecular Biosciences Division, Cardiff School of Biosciences, Cardiff University, Sir Martin Evans Building, Park Place, Cardiff, UK

[6]School of Medicine, Cardiff University, UK

**[*]Corresponding author at:**

Microbiomes, Microbes and Informatics Group, Organisms and Environment Division, School of Biosciences, Cardiff University, Sir Martin Evans Building, Park Place, Cardiff, UK

| **Title** | **Page** |
|---|---|
| **Supplementary Tables and Figures accompanying the main manuscript** | |
| Table S1. Identification of fungi by culture and mycobiota analysis | 3 |
| Table S2. Relative abundance (%) of key genera in BAL1, BAL2, BAL3 and IS samples | 4 |
| Table S3. ASVs identified as significantly associated with sample types by indicator species analysis | 5 |
| Figure S1. Distribution of sequence reads at species level in BAL and IS samples | 6 |
| Figure S2. Microbiota profiles of 25 matched BAL1, BAL2, BAL3 and IS sample sets | 7 |
| Figure S3. Fungal diversity and genus relative abundance associations with increasing age | 8 |
| **Supplementary methods and results** | |
| Quantitative PCR (qPCR) | 9-10 |
| Sequencing controls | 11-15 |
| Sequencing statistics, decontamination and processing | 16-17 |
| Taxonomic classification of fungi | 18 |
| **References** | 19 |
| **Final ASV table and associated metadata (separate Excel document)** | |
| **Bioinformatics scripts and R code (available at: https://github.com/Beky-Weiser/CFSpIT-ITS2-Microbiota-Analysis)** | |

### Table S1. Identification of fungi by culture and mycobiota analysis

| Sample | Culture identification | Mycobiota analysis relative abundance (%)* | | | | | | | | | |
|---|---|---|---|---|---|---|---|---|---|---|---|
| | | Candida | Aspergillus | Dipodascus | Lecanicillium | Simplicillium | Diutina | Exophiala | Penicillium | Wickerhamiella | Thelebolus |
| **CF147BAL1** | Light growth yeast | 93.3 | 1.5 | 1.0 | 1.1 | 0.9 | 0.0 | 0.1 | 0.0 | 0.2 | 0.0 |
| **CF147BAL2** | Light growth yeast and fungus | 93.3 | 0.4 | 0.5 | 2.5 | 1.5 | 0.0 | 0.1 | 0.0 | 0.1 | 0.0 |
| **CF147BAL3** | Moderate growth yeast and light growth fungus | 98.2 | 0.3 | 0.2 | 0.3 | 0.4 | 0.0 | 0.1 | 0.0 | 0.1 | 0.0 |
| **CF147IS** | Light growth yeast | 96.9 | 2.4 | 0.1 | 0.1 | 0.0 | 0.0 | 0.0 | 0.0 | 0.1 | 0.0 |
| **CF152IS** | Candida | 76.7 | 4.0 | 3.9 | 3.0 | 0.5 | 0.0 | 0.0 | 0.1 | 3.2 | 0.0 |
| **CF164IS** | Candida | 92.8 | 1.0 | 0.5 | 0.0 | 0.5 | 0.0 | 0.0 | 0.0 | 0.1 | 0.0 |
| **CF178BAL3** | Light growth yeast | 1.1 | 0.6 | 0.0 | 4.9 | 4.2 | 0.7 | 11.5 | 1.7 | 0.0 | 0.0 |
| **CF181BAL1** | Candida, Aspergillus | 13.1 | 9.9 | 0.1 | 0.1 | 3.0 | 21.2 | 6.0 | 3.7 | 0.0 | 0.0 |
| **CF181BAL2** | Aspergillus, fungus mycellial-like growth | 0.8 | 20.9 | 0.1 | 0.0 | 0.1 | 1.6 | 62.5 | 0.0 | 0.0 | 0.0 |
| **CF181BAL3** | Aspergillus, fungus mycellial-like growth | 0.6 | 96.6 | 0.0 | 0.0 | 0.1 | 1.2 | 0.1 | 0.1 | 0.0 | 0.0 |
| **CF182BAL1** | Candida, Aspergillus | 4.2 | 24.9 | 0.0 | 0.2 | 0.1 | 15.9 | 23.9 | 4.1 | 0.0 | 0.0 |
| **CF182BAL2** | Candida, Aspergillus | 0.3 | 9.2 | 0.0 | 0.3 | 0.2 | 0.8 | 80.1 | 0.4 | 0.5 | 0.0 |
| **CF182BAL3** | Candida, Aspergillus | 2.3 | 11.3 | 0.5 | 44.9 | 17.8 | 0.9 | 0.5 | 0.4 | 3.2 | 0.0 |
| **CF185BAL2** | Aspergillus | 0.2 | 1.9 | 0.0 | 54.0 | 29.8 | 0.3 | 0.0 | 0.0 | 0.0 | 0.0 |
| **CF194IS** | Yeast | 43.5 | 0.2 | 0.1 | 12.5 | 16.1 | 0.2 | 0.1 | 0.1 | 0.0 | 0.0 |
| **CF55IS** | Candida | 51.3 | 8.7 | 4.7 | 5.2 | 7.5 | 0.5 | 0.2 | 1.7 | 0.6 | 0.1 |
| **CF96BAL1** | Candida | 76.9 | 0.3 | 2.8 | 0.2 | 1.8 | 0.0 | 0.0 | 0.0 | 2.7 | 0.0 |
| **CF96BAL2** | Candida | 84.3 | 2.5 | 2.5 | 0.1 | 0.3 | 0.1 | 0.0 | 0.0 | 1.8 | 0.0 |
| **CF96BAL3** | Candida | 1.6 | 5.9 | 0.1 | 0.2 | 0.6 | 0.1 | 0.1 | 1.7 | 2.5 | 0.1 |
| **CF96IS** | Candida | 4.2 | 83.4 | 2.5 | 1.0 | 2.7 | 0.0 | 0.0 | 0.0 | 1.3 | 0.1 |

*Top 10 genera by total abundance across the dataset are shown, cells are coloured from high relative abundance (green) to low relative abundance (white)

**Table S2. Relative abundance (%) of key genera in BAL1, BAL2, BAL3 and IS samples**

| Genus | ALL | | | BAL1 | | | BAL2 | | | BAL3 | | | IS | | |
|---|---|---|---|---|---|---|---|---|---|---|---|---|---|---|---|
| | Average | Min | Max | Average | Min | Max | Average | Min | Max | Average | Min | Max | Average | Min | Max |
| **Candida*** | 14.43 | 0.02 | 98.20 | 12.73 | 0.22 | 93.25 | 9.39 | 0.02 | 93.25 | 9.19 | 0.10 | 98.20 | 26.41 | 0.28 | 96.88 |
| **Aspergillus** | 13.42 | 0.07 | 96.58 | 12.14 | 0.15 | 68.48 | 14.20 | 0.40 | 85.72 | 12.33 | 0.07 | 96.58 | 15.01 | 0.18 | 83.37 |
| **Dipodascus** | 7.14 | 0.00 | 54.30 | 8.64 | 0.00 | 48.93 | 6.75 | 0.00 | 54.30 | 4.48 | 0.00 | 28.83 | 8.69 | 0.00 | 48.75 |
| **Lecanicillium** | 6.82 | 0.00 | 53.95 | 6.68 | 0.03 | 34.08 | 7.26 | 0.03 | 53.95 | 9.32 | 0.03 | 44.92 | 4.02 | 0.00 | 24.15 |
| **Simplicillium** | 6.45 | 0.02 | 47.72 | 6.32 | 0.02 | 47.72 | 7.47 | 0.03 | 40.12 | 8.12 | 0.05 | 45.13 | 3.87 | 0.02 | 23.82 |
| **Exophiala** | 3.16 | 0.00 | 80.05 | 1.38 | 0.00 | 23.92 | 7.22 | 0.00 | 80.05 | 0.87 | 0.00 | 11.52 | 3.18 | 0.00 | 50.52 |
| **Lomentospora** | 0.84 | 0.00 | 68.82 | 0.21 | 0.00 | 2.60 | 0.11 | 0.00 | 1.53 | 3.00 | 0.00 | 68.82 | 0.04 | 0.00 | 0.20 |
| **Scedosporium** | 0.03 | 0.00 | 1.20 | 0.01 | 0.00 | 0.10 | 0.10 | 0.00 | 1.20 | 0.02 | 0.00 | 0.37 | 0.01 | 0.00 | 0.08 |

*IS relative abundance was significantly increased compared to BAL1 (p=0.03311), BAL2 (p=0.00113 and BAL3 (p=0.00551)
Cells are coloured from high relative abundance (green) to low relative abundance (white)

**Table S3. ASVs identified as significantly associated with sample types by indicator species analysis**

| ASV | Stat | p-value | Significance level |
|---|---|---|---|
| **BAL1** | | | |
| Candida_krusei_ASV29 | 0.314 | 0.0076 | ** |
| Order_Saccharomycetales_ASV60 | 0.273 | 0.0287 | * |
| Chrysosporium_lobatum_ASV398 | 0.255 | 0.0318 | * |
| **BAL3** | | | |
| Kingdom_Fungi_ASV589 | 0.368 | 0.0029 | ** |
| Curvularia_lunata_ASV198 | 0.305 | 0.0008 | *** |
| Order_Sebacinales_ASV120 | 0.295 | 0.0208 | * |
| Genus_Geminibasidium_ASV95 | 0.278 | 0.0143 | * |
| Curvularia_intermedia_ASV81 | 0.274 | 0.0431 | * |
| Genus_Periconia_ASV649 | 0.27 | 0.0316 | * |
| Kingdom_Fungi_ASV127 | 0.261 | 0.0435 | * |
| Order_Sebacinales_ASV332 | 0.261 | 0.0115 | * |
| Saitozyma_podzolica_ASV28 | 0.26 | 0.0437 | * |
| Genus_Geminibasidium_ASV96 | 0.26 | 0.0491 | * |
| Kingdom_Fungi_ASV227 | 0.254 | 0.0303 | * |
| Genus_Simplicillium_ASV210 | 0.239 | 0.0429 | * |
| Genus_Geminibasidium_ASV351 | 0.223 | 0.0025 | ** |
| Cladosporium_cladosporioides_ASV38 | 0.217 | 0.0411 | * |
| **IS** | | | |
| Saccharomyces_cerevisiae_ASV74 | 0.325 | 0.0029 | ** |
| Kingdom_Fungi_ASV53 | 0.322 | 0.0009 | *** |
| Candida_albicans_ASV10 | 0.281 | 0.0207 | * |
| Phylum_Rozellomycota_ASV149 | 0.268 | 0.0294 | * |
| Candida_albicans_ASV14 | 0.266 | 0.0274 | * |
| Genus_Trichoderma_ASV324 | 0.177 | 0.0116 | * |
| Chordomyces_antarcticus_ASV280 | 0.177 | 0.0135 | * |
| **BAL1 + BAL3** | | | |
| Cladosporium_cladosporioides_ASV101 | 0.266 | 0.0307 | * |

Rows with ASVs within the Top 50 most abundant ASVs are shaded orange; Significance levels, p<0.05 = *, p<0.01 = **, p<0.001 = ***

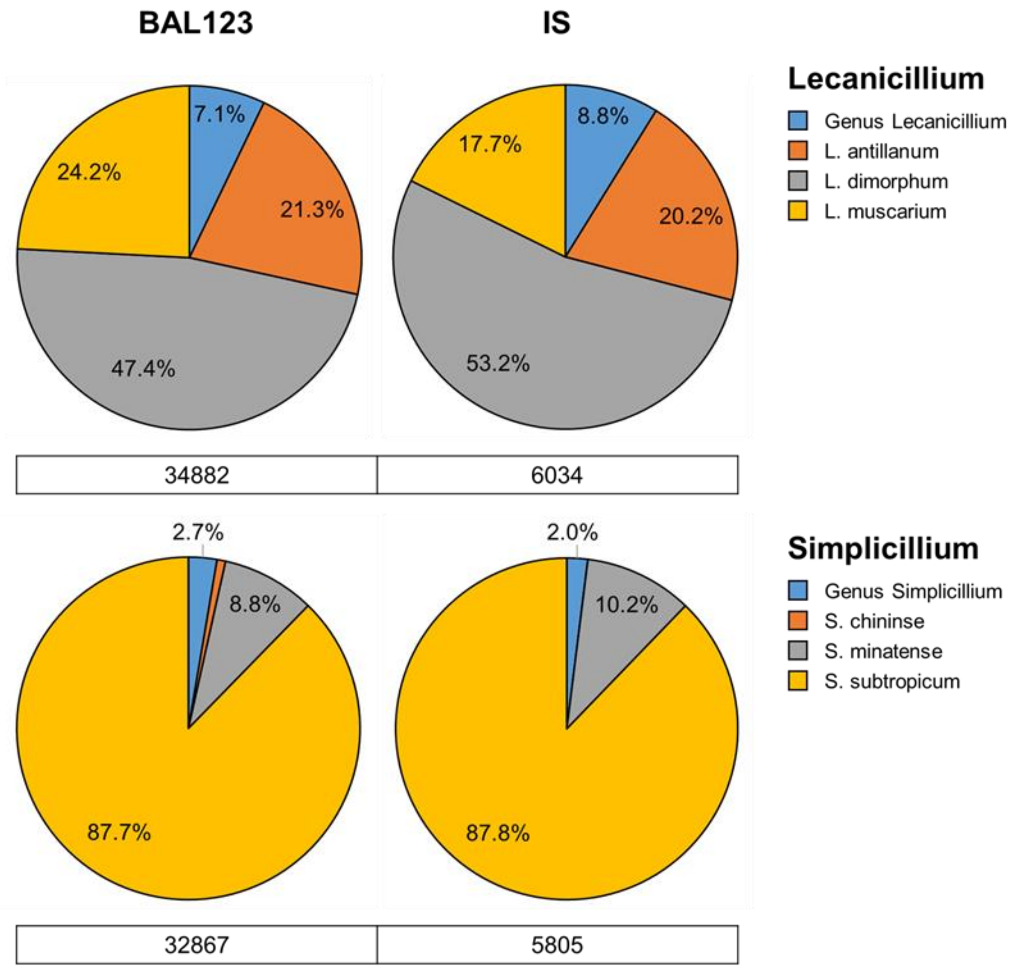


**Figure S1. Distribution of sequence reads at species level in BAL and IS samples.** Two of the most abundant and prevalent genera (*Lecanicillium* and *Simplicillium*) are shown. Reads from BAL1, BAL2 and BAL3 samples are consolidated into one group as 'BAL123'. Pie charts display the proportion of sequence reads assigned to different species within a genus. Total sequence reads for each genus are below each pie chart. Other abundant and prevalent genera (*Dipodascus* and *Diutina*) and presumptive cystic fibrosis pathogens (*Lomentospora* and *Scedosporium*) are not shown as sequence reads were assigned to a single species within these genera (*Dipodascus geotrichum*, *Diutina catenulata*, *Lomentospora prolificans* and *Scedosporium boydii*).

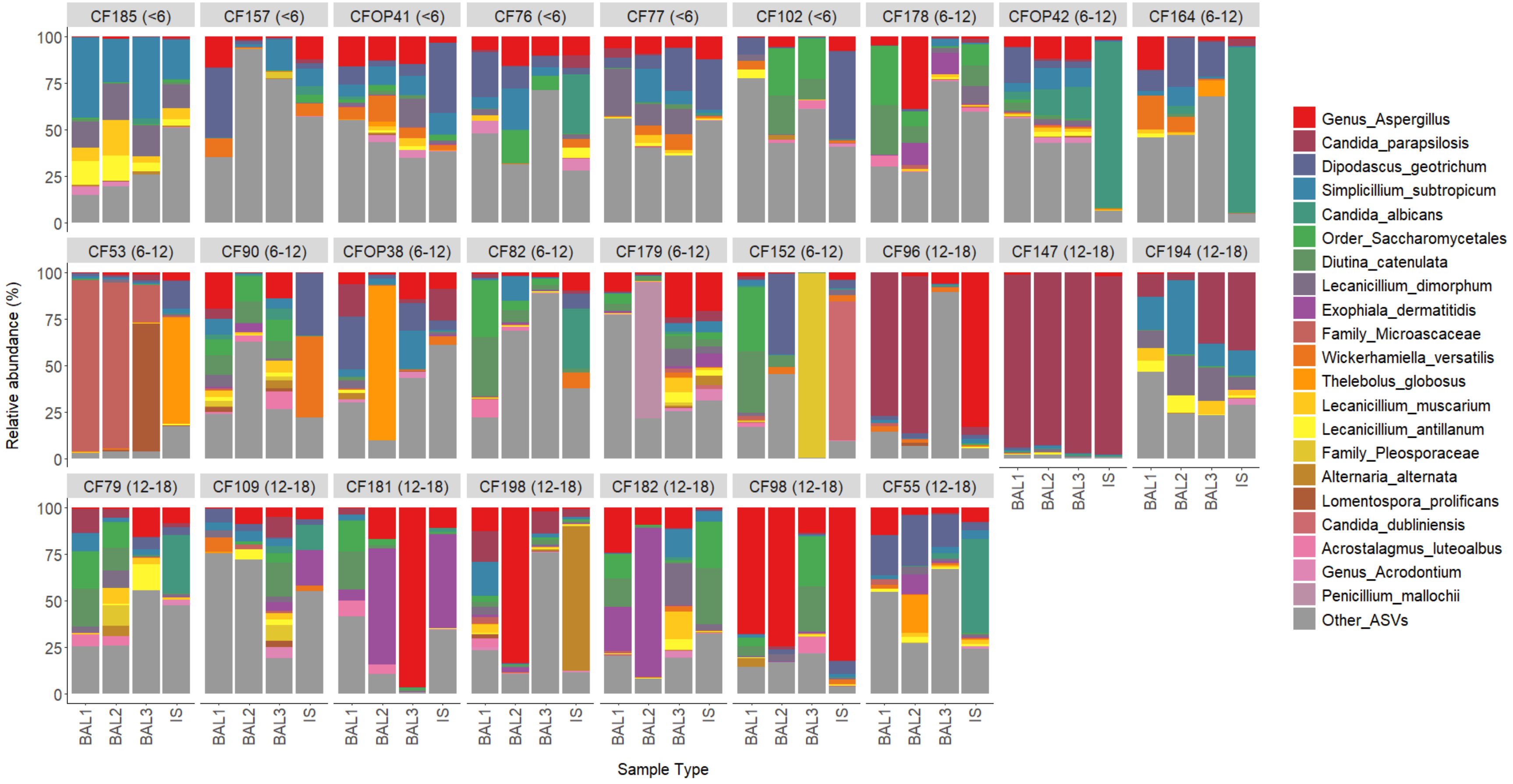


**Figure S2. Microbiota profiles of 25 matched BAL1, BAL2, BAL3 and IS sample sets.** Stacked bar charts show the relative abundance of the top 25 most abundant ASVs consolidated to lowest taxonomic rank and all other ASVs consolidated into a single group. Samples are grouped by set, patient age group is displayed in brackets to the right of the sample set number and the taxa colour key is displayed at the right of the figure. Two patients provided sample sets on two separate occasions (CF96 and CF198; CFOP38 and CF194).

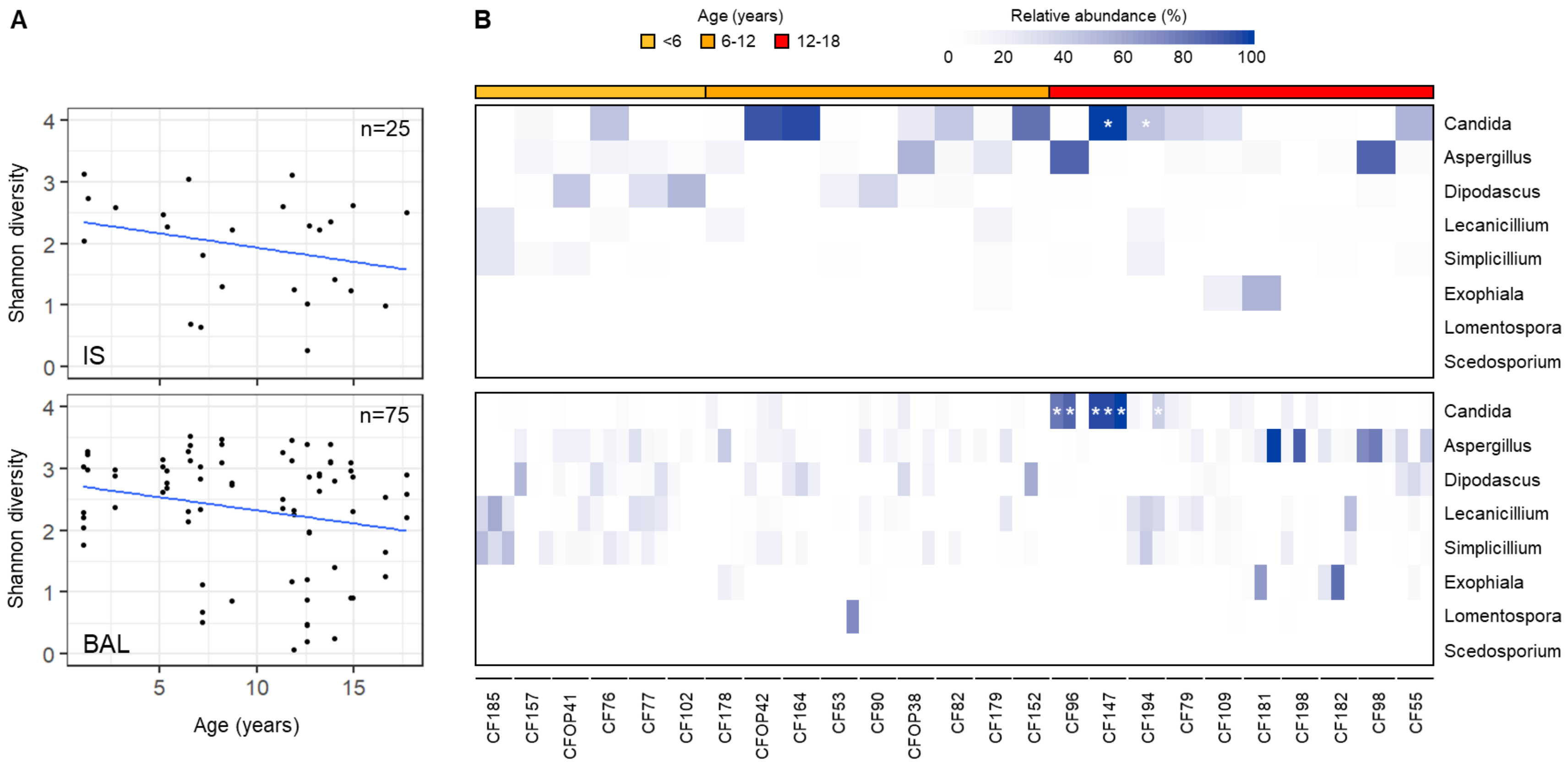


**Figure S3. Fungal diversity and genus relative abundance associations with increasing age.** Scatterplots and linear regression **(A)** were used to examine the relationship between Shannon diversity and age for IS samples (n=25; top panel) and BAL samples (n=75; bottom panel). The regression lines show a trend for decreased diversity with increasing age although this was not significant. Heatmaps **(B)** show the relative abundance of the five most abundant genera across the dataset and three presumptive cystic fibrosis pathogens for IS samples (n=25; top panel) and BAL samples (n=75; bottom panel). Samples are arranged in order of increasing age and the sample set number is given at the bottom of the figure. All three BAL samples are presented for each set in the order BAL1-BAL2-BAL3. The colour key for age and relative abundance is displayed at the top of the figure. For *Candida*, cells marked with a white asterisk have >35% relative abundance attributed to *C. parapsilosis*.

**Supplementary methods and results**

### 1. Quantitative PCR (qPCR)

Quantification of total fungal load was performed using a TaqMan® qPCR assay targeting the fungal 18S rRNA gene as described by Liu *et al.* (2012)(1). The qPCR assay was performed on 24/25 samples sets; CFOP42 could not be tested as there was not sufficient DNA left after ITS2 region sequencing. Reaction volumes were 10 µl and comprised 1X Platinum qPCR Supermix-UDG with ROX (Invitrogen), 1.8 µM forward and reverse primers (Supplementary Table S4; Eurofins Genomics), 225 nM TaqMan® probe (Thermofisher Scientific), 1 µl template DNA and nuclease free water (Severn Biotech Ltd.). Reactions were performed in triplicate alongside template-free negative controls and a qPCR standard dilution series. The qPCR standard was a 951 bp region of the 18S rRNA gene that encompassed the qPCR target. This region was amplified from *Candida albicans* SC 5314 and tenfold dilutions ($10^8$-$10^2$ copies) made in nuclease free water for use in the qPCR (primers in Supplementary Table S4). Accurate and consistent loading of 384-well qPCR plates was achieved through automated pipetting with an Opentrons OT-2 robot. The QuantStudio 7 Flex Real-Time PCR System (Applied Biosystems, Thermofisher Scientific) was used with the following qPCR cycling conditions: UNG treatment at 50 °C for 3 min, Taq activation at 95 °C for 10 min, followed by 50 cycles of denaturation at 95 °C for 15 s, annealing and extension at 65°C for 1 min and a plate read. Quality control of qPCR results within a run was performed as described previously (2). Two qPCR runs were performed to obtain two biological replicates per sample which were averaged to give the final result (Supplementary Table S5). $R^2$ values were 0.999 for both runs and efficiency values were 84.9% for run 1 and 87.2% for run 2. 18S rRNA gene copy number was consistently low; only 13/100 samples had over 100 copies/µl, only 3 of which had over 1000 copies/µl (linked to >95% relative abundance of *C. parapsilosis*). Although previous studies have reported that this qPCR assay has poor sensitivity (3) and different regions were targeted in qPCR (18S) and sequencing (ITS2), such low copy numbers were unexpected and in contrast to the high number of reads obtained from ITS2 region sequencing (see section 3). We wanted to report this result for study transparency and to demonstrate that qPCR and read data do not always correlate. Subsequent sections demonstrate how we performed quality control on our sequencing read data (without qPCR data) and reassuringly found mycobiota patterns replicated in multiple samples from the same individual (Figure S2), suggesting a genuine biological signal.

**Table S4. PCR and qPCR primers**

| PCR target | Primers/ Probe | Sequence 5' > 3' | Annealing temp (°C) | Product (bp)* | Reference |
|---|---|---|---|---|---|
| qPCR standard (18S) | Forward | TGGCGAACCAGGACTTTTAC | 56 | 951 | This study |
| | Reverse | AGGCCTCACTAAGCCATTCA | | | |
| qPCR target (18S) | Forward | 5′-GG**R**AAACTCACCAGGTCCAG-3′ | 65 | 353 | Liu et al. (2012) (1) |
| | Reverse | 5′-G**SW**CTATCCCCA**K**CACGA-3′ | | | |
| | Probe | 6FAM-5′-TGGTGCATGGCCGTT-3′-MGBNFQ | | | |

*Product size for *C. albicans* SC 5314

**Table S5. 18S rRNA gene copies per ul of DNA extracted from each sample as determined by qPCR**

| Sample | BAL1 | BAL2 | BAL3 | IS |
|---|---|---|---|---|
| **CF102** | 7 | 0 | 0 | 1 |
| **CF109** | 56 | 0 | 0 | 40 |
| **CF147** | 721 | 3843 | 15614 | 15829 |
| **CF152** | 0 | 0 | 0 | 67 |
| **CF157** | 0 | 0 | 0 | 40 |
| **CF164** | 0 | 11 | 1 | 157 |
| **CF178** | 0 | 7 | 59 | 4 |
| **CF179** | 3 | 3 | 0 | 0 |
| **CF181** | 8 | 70 | 125 | 95 |
| **CF182** | 19 | 111 | 8 | 0 |
| **CF185** | 34 | 5 | 8 | 8 |
| **CF194** | 33 | 0 | 89 | 110 |
| **CF198** | 1 | 0 | 19 | 7 |
| **CF53** | 156 | 358 | 484 | 0 |
| **CF55** | 0 | 0 | 0 | 205 |
| **CF76** | 1 | 5 | 0 | 10 |
| **CF77** | 0 | 2 | 22 | 0 |
| **CF79** | 0 | 0 | 0 | 60 |
| **CF82** | 0 | 0 | 4 | 17 |
| **CF90** | 0 | 0 | 4 | 0 |
| **CF96** | 86 | 62 | 17 | 65 |
| **CF98** | 37 | 25 | 20 | 104 |
| **CFOP38** | 49 | 0 | 24 | 48 |
| **CFOP41** | 2 | 0 | 0 | 5 |

Samples with between 100 and 1000 copies are shaded blue, samples with over 1000 copies are shaded green. Blank nuclease free water controls were also included and there was no amplification in these.

### 2. Sequencing controls

#### 2.1 Mock community

A fungal mock community (Mycobiome Genomic DNA Mix, MSA-1010; ATCC) was included in the sequencing run to investigate mycobiome profiling accuracy. The expected mycobiome profile of the mock community was compared to the mycobiome profile obtained after sequencing and QIIME2-UNITE database analysis with and without read filtering and subsampling (Table S6; visual representation in Figure S4). For both unprocessed and processed reads, 7 out of the 10 fungal species in the mock community were correctly identified at species level, with the remaining 3 correctly identified to genus level (Table S6). The top 10 most abundant ASVs in the sequenced mock communities also matched the 10 expected mock community species and the remaining ASVs represented less than 1.3% of the total relative abundance. However, there was deviation from the expected relative abundance of 10% for each species, with certain species reaching almost triple the expected relative abundance (*Fusarium keratoplasticum*, 28.28%) and others being almost 100-fold lower (*Malassezia globose*, 0.38%). Therefore, although sequencing and analysis was able to correctly identify fungal taxa, at least to genus level, there may have been instances where the abundances of certain taxa were under- or over-estimated. This is a known limitation of microbiota analysis (4) that we could unfortunately not correct for in this study. The read filtering and subsampling quality control steps (section 3) reduced the relative abundances of non-mock community taxa in the mock community control (Table S6; 1.28% to 0.9%) and made the sequencing dataset as robust as possible for mycobiota analysis.

**Table S6. Relative abundance of fungal species in the mock community sequencing control**

| Fungal species | Relative abundance (%) | | |
|---|---|---|---|
| | Expected | No filtering or subsampling (105221 reads) | ASVs <0.001% total read abundance removed & subsampled to 6000 reads |
| Fusarium keratoplasticum | 10 | 28.18 | 28.28 |
| Penicillium (chrysogenum)* | 10 | 19.25 | 19.65 |
| Candida albicans | 10 | 9.39 | 8.88 |
| Cryptococcus neoformans | 10 | 9.90 | 10.15 |
| Trichophyton (interdigitale)** | 10 | 6.38 | 6.75 |
| Aspergillus (fumigatus)*** | 10 | 4.81 | 5.10 |
| Candida glabrata | 10 | 2.74 | 2.98 |
| Malassezia globose | 10 | 0.38 | 0.30 |
| Saccharomyces cerevisiae | 10 | 4.88 | 4.60 |
| Cutaneotrichosporon dermatis | 10 | 12.82 | 12.40 |
| Others (consolidated) | 0 | 1.28 | 0.90 |

*Penicillium chrysogenum in the mock community was identified as Genus_Pencillium
**Trichophyton interdigitale in the mock community was identified as Trichophyton mentagrophytes
*** Aspergillus fumigatus in the mock community was identified as Genus_Aspergillus

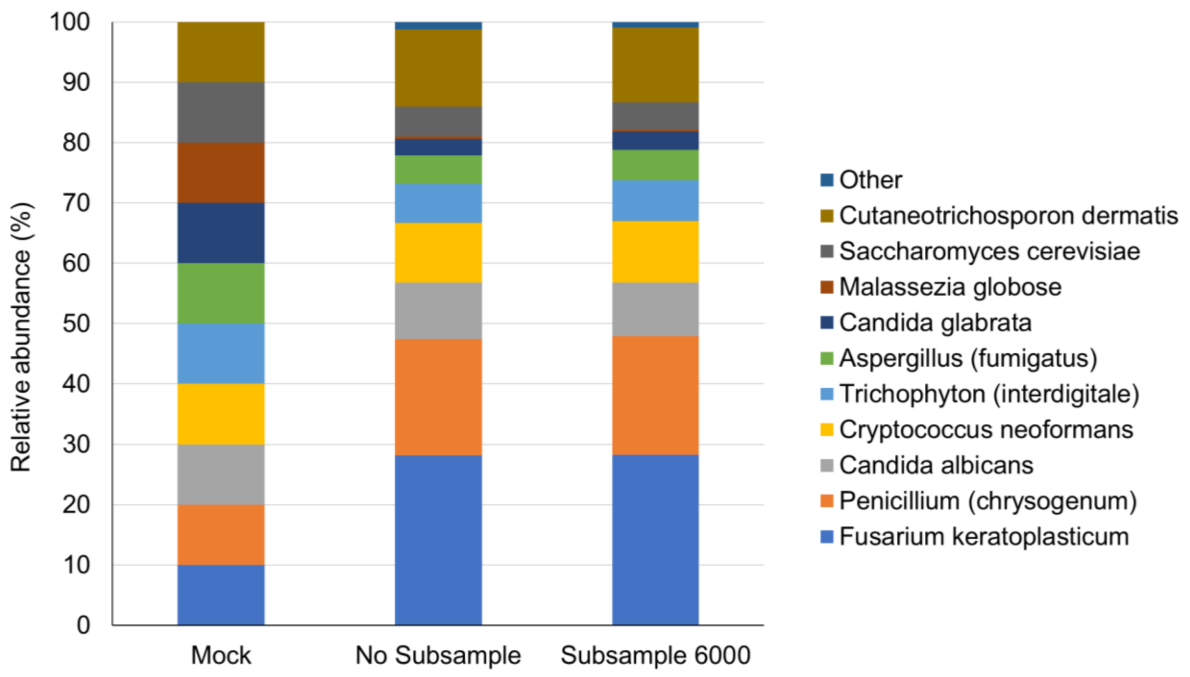


**Figure S4. Fungal diversity and abundance of the mock community MSA-1010 and the mock community results after sequencing.** Results are shown with and without read filtering and subsampling. The key indicates the species in the samples.

#### 2.2 DNA extraction controls

Three DNA extraction controls taken from three different batches of DNA extraction kits were included in the study. The numbers of sequencing reads obtained for the blanks were highly variable (Blank 1=123627, Blank 2=5971 and Blank 3=5897) and the overall mycobiota profiles were dissimilar in terms of diversity and taxa (Figure S5). Blank 1 was less diverse than Blank 2 and Blank 3, with lower overall numbers of ASVs (Blank 1=61, Blank 2=153, Blank 3=100). Blank 1 contained a dominant ASV (the fungal plant pathogen *Choanephora cucurbitarum*) comprising 94% relative abundance. Blank 2 and Blank 3 did not have a dominant species, and their most abundant ASVs reached 10% (*Diutina catenulata*) and 31% (*Dipodascus geotrichum*), respectively. Whilst certain ASVs were shared between the top 5 most abundant ASVs of the three blanks (Table S7), no consistent contaminant pattern could be identified. As there was almost no overlap between the predominant ASV in Blank 1 and samples in the study (only 4/100 samples carried the ASV and at <2.2% sequence reads) it was unlikely to pose a problem as a kit contaminant. However, ASVs including *Diutina catenulata* (present in 88/100 study samples at up to 32% sequence reads) and *Dipodascus geotrichum* (present in 93/100 study samples at up to 42% sequence reads) did overlap between blanks and study samples indicating that sequence decontamination should be investigated (see section 3).

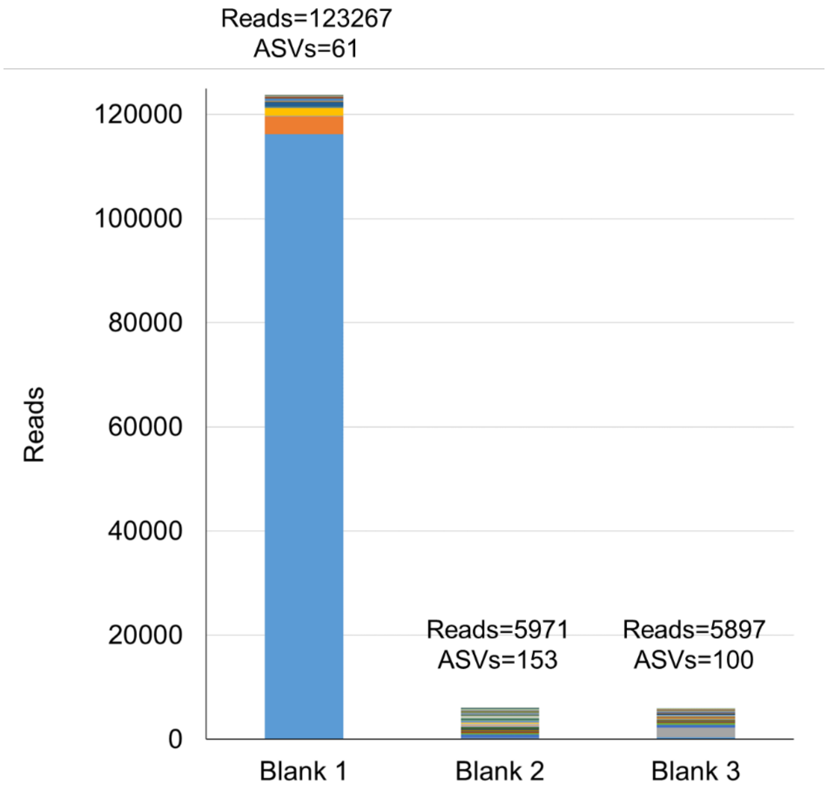


**Figure S5. Fungal community profiles observed in the DNA extraction controls.** Read numbers and total ASVs numbers in each blank is shown above each stacked bar. No colour key is given to identify the different ASVs in the stacked bar charts as over 229 ASVs were identified across the three blanks. The predominant ASV in Blank 1 was *Choanephora cucurbitarum* (94% relative abundance), in Blank 2 was *Diutina catenulate* (10% relative abundance) and in Blank 3 was *Dipodascus geotrichum* (31% relative abundance).

**Table S7. Top 5 most abundant ASVs in the DNA extraction blanks**

| ASV taxon ID | Relative abundance (%) |
|---|---|
| **Blank 1** | |
| Choanephora_cucurbitarum | 94.01 |
| Bipolaris_yamadae | 2.73 |
| Dipodascus_geotrichum | 1.23 |
| Choanephora_cucurbitarum | 0.61 |
| Diutina_catenulata | 0.16 |
| **Blank 2** | |
| Diutina_catenulata | 10.13 |
| Phialemoniopsis_curvata | 5.78 |
| Order_Saccharomycetales | 5.54 |
| Genus_Metschnikowia | 4.62 |
| Trametes_trogii | 4.61 |
| **Blank 3** | |
| Dipodascus_geotrichum | 31.47 |
| Diutina_catenulata | 8.21 |
| Choanephora_cucurbitarum | 7.19 |
| Order_Saccharomycetales | 7.00 |
| Order_Russulales | 3.34 |

Blanks sharing the same species belonging to the same ASVs are coloured. Blanks sharing the same species belonging to different ASVs are not coloured.

### 3. Sequencing statistics, decontamination and processing

In total, 821385 reads were obtained from the 100 samples making up 25 matched sets. Broken down by sample type, the average read numbers were: BAL, 84692; BAL2, 85707; BAL3, 84265, and; IS, 75391 (Table S8). Read numbers were consistently high (≥11,000), in contrast to the qPCR results based on 18S rRNA copy number which suggested that nearly all of the samples were low biomass.

As low biomass samples are prone to sequence contaminants from kits and reagents (5) and there were ASVs that overlapped between the DNA extraction controls and study samples, we wanted to investigate the possibility that contamination had occurred. The R package Decontam (5) has been widely used for this purpose and uses both ‘frequency’ (DNA concentration/qPCR and read data) and ‘prevalence’ (read data only) methods to compare ASVs in blank controls and study samples to identify potential contaminants. As our qPCR data suggested that 35 out of 100 samples had no fungal DNA (even though read numbers were high) we decided that the prevalence method was not suitable and used only the frequency method. The frequency method identified 103 ASVs as potential contaminants and although it did identify the predominant contaminant in Blank 1 (Choanephora_cucurbitarum), it also identified 7/10 ASVs of the mock community as contaminants. We decided that these results were inconclusive and devised our own strategy to process the reads and minimise potential contamination from kits and reagents:

1. **Filter out low abundance ASVs from the dataset.** As there is no consensus filtering threshold between different mycobiota studies, a range of different thresholds were considered (removing ASVs representing less than 1/0.1/0.02/0.01% of the total number of sequence reads). Ultimately, we chose to remove ASVs representing less than 0.01% of the total number of sequence reads, which greatly reduced the number of ASVs (5891 to 680) but retained 94.37% of the reads.
2. **Subsample the sequence reads.** We chose to subsample to 6000 reads as this was just above the number of reads in Blank 2 and Blank 3 and below all study samples. Blank 1 had a higher read number but was not taken into consideration as its predominant contaminant was at very low frequency and abundance in study samples so was a low contamination risk. After subsampling, sample coverage was calculated at ≥99.5%.

Comparison of the mock community before and after ASV filtering and sequence subsampling indicated a high level of concordance between the relative abundances of mock community species and a reduction in the relative abundances of non-mock community species (Table S6, Figure S4). We were confident that these quality control measures produced the best possible dataset for mycobiota analysis.

**Table S8. ITS2 region sequence read numbers for BAL1, BAL2, BAL3 and IS samples**

| Sample | BAL1 | BAL2 | BAL3 | IS |
|---|---|---|---|---|
| CF102 | 104150 | 71457 | 92743 | 84794 |
| CF109 | 88135 | 94606 | 78149 | 50871 |
| CF147 | 33439 | 112732 | 124051 | 103244 |
| CF152 | 98659 | 115467 | 92950 | 57313 |
| CF157 | 114782 | 71586 | 102215 | 75548 |
| CF164 | 85763 | 97769 | 34107 | 128413 |
| CF178 | 95926 | 65067 | 72061 | 59845 |
| CF179 | 72994 | 54197 | 79339 | 74653 |
| CF181 | 93150 | 78180 | 90655 | 27230 |
| CF182 | 112234 | 89936 | 82528 | 53285 |
| CF185 | 83016 | 81785 | 108026 | 69387 |
| CF194 | 82179 | 102269 | 82237 | 76657 |
| CF198 | 65332 | 61952 | 59155 | 64680 |
| CF53 | 131500 | 128726 | 127422 | 119685 |
| CF55 | 116342 | 109953 | 114551 | 97371 |
| CF76 | 99952 | 42621 | 98021 | 71690 |
| CF77 | 90450 | 111913 | 131847 | 17858 |
| CF79 | 26808 | 33642 | 40422 | 95437 |
| CF82 | 26084 | 11000 | 57024 | 82553 |
| CF90 | 46182 | 73701 | 52667 | 17584 |
| CF96 | 129926 | 100340 | 62515 | 129603 |
| CF98 | 72716 | 116209 | 29012 | 39599 |
| CFOP38 | 79996 | 113217 | 89484 | 108211 |
| CFOP41 | 75287 | 99634 | 89496 | 65202 |
| CFOP42 | 92302 | 104725 | 115936 | 114071 |
| **Read statistics** | | | | |
| Minimum | 26084 | 11000 | 29012 | 17584 |
| Maximum | 131500 | 128726 | 131847 | 129603 |
| Average | 84692.16 | 85707.36 | 84264.52 | 75391.36 |

### 4. Taxonomic classification of fungi

All taxonomic classifications made by the QIIME2-UNITE database pipeline were adhered to except for 2 ASVs identified as *Issatchenkia orientalis* (one ASV) and *Scedosporium prolificans* (one ASV). *I. orientalis* was recently shown to be genetically indistinct from *Candida krusei*, *Candida glycerinogenes* and *Pichia kudriavzevii* (6). In this study, *I. orientalis* was changed to *C. krusei* to reflect its clinical relevance; *C. krusei* is responsible for approximately 2% of human *Candida* infections, whereas the other three species are largely associated with the natural environment and biotechnological applications (6). In addition, *S. prolificans* was changed to *Lomentospora prolificans* to reflect the taxonomic revision of 2014, which found that *S. prolificans* was genetically distinct from other *Scedosporium* species (7).

Surprisingly, although *A. fumigatus* is one of the most common fungi isolated from CF respiratory samples, none of the ASVs identified as *Aspergillus* were further classified to *A. fumigatus* by the QIIME2-UNITE database pipeline. Of the 34 Aspergillus ASVs, 13 were classified to species level and 21 were classified only to genus level. When the sequences of the 21 Genus_Aspergillus ASVs were analysed using the NCBI blastn tool (https://blast.ncbi.nlm.nih.gov/Blast.cgi; Internal transcribed spacer region (ITS) from Fungi type and reference material database) 4 ASVs comprising 39765/70443 (56%) of the total Genus_Aspergillus sequencing reads were identified as *A. fumigatus* (coverage: 100%, percentage identity: ≥99.46, e-value ≤2.00E-92). One of these ASVs (ASV_2) was the second most prevalent ASV across the entire dataset and was also identified at 5.1% relative abundance in the mock community, which contained 10% *A. fumigatus*. We therefore conclude that detection of *A. fumigatus* was low in the dataset, not because it wasn't present, but because of difficulties in fungal ASV classification.
